# Epidemiological aspects of COVID-19 disease in India during nationwide lockdown phase-An empirical data-based analysis and its implications on interrupting the transmission

**DOI:** 10.1101/2020.07.16.20155903

**Authors:** Vanamail Perumal

## Abstract

**Background:** Covid-19 disease is pandemic in more than 85% of the countries in the world, with about 10 million cases and 0.5 million deaths as on July 2, 2020. In India reporting of the first case was on January 30, 2020, and to prevent rapid community spread of the disease nationwide lockdown phase was imposed from March 25- June 1, 2020. Our objective was to assess various epidemiological measures during the lockdown phase.

**Methods:** We used daily reporting of confirmed cases by the Ministry of Health and Family Welfare, Government of India during the period March 19-June 1, 2020. Using statistical packages STATA and R-packages, we fitted three statistical distributions (Gamma, Weibull and Log-normal) to the daily recorded new cases. We estimated daily incidence rate and death rate per million population, generation time and Basic Reproduction numbers.

**Results:** During the lockdown phase, the daily per cent increase in the cumulative number of cases showed negative exponential growth with 0.022 as an instantaneous rate of decrease. The average incidence rate with a 95% confidence interval (CI) was 1.84 (1.43-2.25). Day specific incidence rate per million (revealed the exponential pattern with 0.069 as the instantaneous rate of increase per day, which accounted for the doubling time of the disease (10 days; 95% CI: 9.25-10.93). Case fatality rate (2.92%; 95% CI: 2.82% −3.02%) and overall death rate was 1.14 (95% CI: 0.87-1.41) per million. were abysmally low. Statistical distribution fitting of new cases found to be satisfactory with Gamma distribution. Basic reproduction numbers 1.83 (95% CI: 1.82-1.83) was less.

**Conclusion:** In India, with a population density of about 450 per Km^2^, the virulent of COVID-19 transmission was interrupted significantly with 70 days lockdown during the early transmission stage. We observed a considerable decline in all the epidemiological indices compared to the corresponding indices recorded during the same period in the severely affected countries.

## Introduction

Currently, about 215 out of 251 countries recognized by the United Nations are experiencing with an infectious disease called coronavirus or COVID-19 [1]. Initially, the virus (thought to have transmitted from animals to humans) was detected in the Wuhan city, China, during the last week of December 2019 [2,3]. Since this virulent disease has broken out simultaneously all over the world, Considering the severity and rapid spread of this disease, the World Health Organization (WHO) declared a pandemic. According to COVID-19 Situation Report (No.164) released by the WHO [1] dated July 2, 2020, there were about 10.5 million infected persons and 0.5 million deaths (4.8%). Reporting of the first COVID-19 case in India was on January 30, 2020, originating from China. Steadily increasing and as of July 2, 2020, the Ministry of Health and Family Welfare (MoHFW), Government of India (GOI) have confirmed a total of 6,04,641 cases, 19,148 new cases and 17,834 deaths resulting from the case fatality rate 2.95% in the country [1]. India being the second most populated country, the GOI had expected many COVID-19 cases and started taking various integrated approaches such as screening, treating and quarantining the suspected individual to reduce the disease within a manageable level [4]. While the disease load crossed 500 in the third week of March 2020, considering the value of human life and by compromising the Indian economy, the GOI took an innovative approach called “Nationwide lockdown” for 70 days in four phases (March 25-April 14, April 15-May 3, May 4 – May 17 and May 18-June 1, 2020). During that period all kinds of transports, social gatherings in public places were banned with strict enforcement of law and order. It was ensured that only the essential services were available during that period.

The present study aimed to assess the necessary epidemiological measures of the COVID-19 disease in India during the lockdown phase. It is a well-known fact that India is contributing about 18% of the total world population and ranks number two in the list of countries by population. Therefore, population-based epidemiological measures are vital to compare the severity of the disease with the other affected countries. Empirical based data analysis will have a further implication on preventing the social spread of this disease.

## Materials and Methods

### Data source

National level daily reporting (between 8 AM and 10 AM) of confirmed cases of COVID-19, recovered and deaths are available either in the website of MoHFW, GOI or in COVID-19 situation Reports published by WHO daily. Though reporting the first case was on January 30, 2020, rapid daily increase was noted from March 18, 2020, onwards. In expecting a more significant number of cases by social spread, the GOI had imposed nationwide lockdown period from March 25 to June 1, 2020. For the present analysis, we considered the cumulative number of cases recorded on March 18 as index cases for COVID- 19 transmission in India. Therefore, to assess various epidemiological indices, daily recorded new cases between 8 AM and 10 AM for a period 75 days (March 19, 2020 – June 1, 2020) formed the database for the present analysis. The entire database is available both in MoHFW, GOI website [4] as well as COVID-19 situation reports of WHO [1]. Based on the daily news, we created the essential data in the Excel spreadsheet for the present analysis. We considered midyear projected population of India (∼ 1366 million) in the year 2019 [5] to be susceptible to COVID-19 and assumed to be stationary during the study period.

### Statistical analysis

We performed statistical analysis using Statistical Softwares STATA version 16.0 (College Station, TX: StataCorp LLC. StataCorp) and R0-package in R-version 4.0. We derived the following epidemiological indices;

Daily increase rate (%) in cumulative no. of cases;

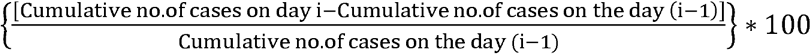

Daily incidence rate per 10 lakh population;

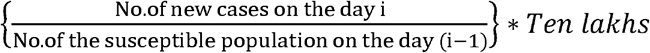

Daily case fatality rate (%)’;

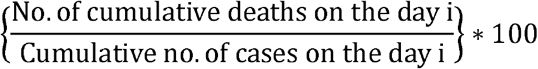

The daily death rate per 10 lakh population;

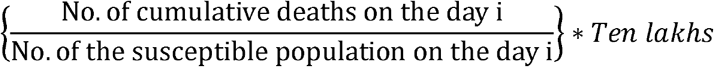

To find out the underlying distribution of day-specific new cases, we fitted popular discrete distributions such as Gamma, Weibull and Log-normal using maximum likelihood estimation method by applying an R-package (“fitdistrplus”). We tested the adequacy of the fitted distribution by comparing goodness of fit statistics (Kolmogorov-Smirnov statistics, Cramer-von Mises statistics and Anderson-Darling statistics) and goodness of fit criteria (Akaike Information Criteria (AIC) and Bayesian Information Criterion (BIC)). Among the three distributions fitted, we selected the distribution that yielded the lowest goodness of fit statistics and goodness of fit criteria as the best one. Further, for the selected best distribution we confirmed the consistency of parameters (shape and scale) estimation using different methods such as matching moment estimation (MME), maximum spacing estimation (MSE) and maximum goodness of fit estimation (MGE).

Generation Time (GT), which describes “the time that elapses between onset of symptoms in the primary case and onset of symptoms of the secondary case” is a crucial epidemiological parameter that describes the of the disease transmission. Therefore, to get the best estimate of GT, we assumed different underlying distributions such as Empirical, Gamma, Weibull and Log-normal. We obtained mean and standard deviation of GT for each distribution by applying R-packages “MASS” and “R0”.

Basic reproduction rate (R0) suggests the transmission potential of a disease. It is the average number of secondary infections produced by an infected case in a susceptible population. Therefore, to know the transmission potential of the COVID-19 disease in India and particularly during different phases of the lockdown period, we estimated R0 with 95% confidence limits using the maximum likelihood estimation and exponential growth method.

Doubling time, the period needed for the number of cases in the endemic to double is another critical sign to assess the severity of the disease. Therefore, using the exponential growth (r), we calculated the doubling time as follows;

Doubling time =ln(2) /*r*

## Results

During the 75 days study period (March 19, 2020-June 1, 2020) day-specific number of new cases increased from 14 on March 19 to 8392 on June 1. The cumulative number of cases were 151 and 1, 90, 535. As on June 1, the total cases comprised of 49% (93322) active cases, 2.8% (5394) deaths and 48.2% (91819) cured cases.

### Daily increasing (%) pattern

The percent increase in the cumulative no. of cases is shown in Fig. 1. We showed a negative exponential pattern between the time (t) and the percent increase (y). The fitted equation *y* = 19.9 * exp(−0.022*t*) was satisfactory with R^2^-adjusted value 0.73 (P<0.001), AIC (30.1) and RMSE (0.29). At the end of the lockdown phase (June 1, 2020) the predicted percent increase was about 3.8.

**Fig. 1.**
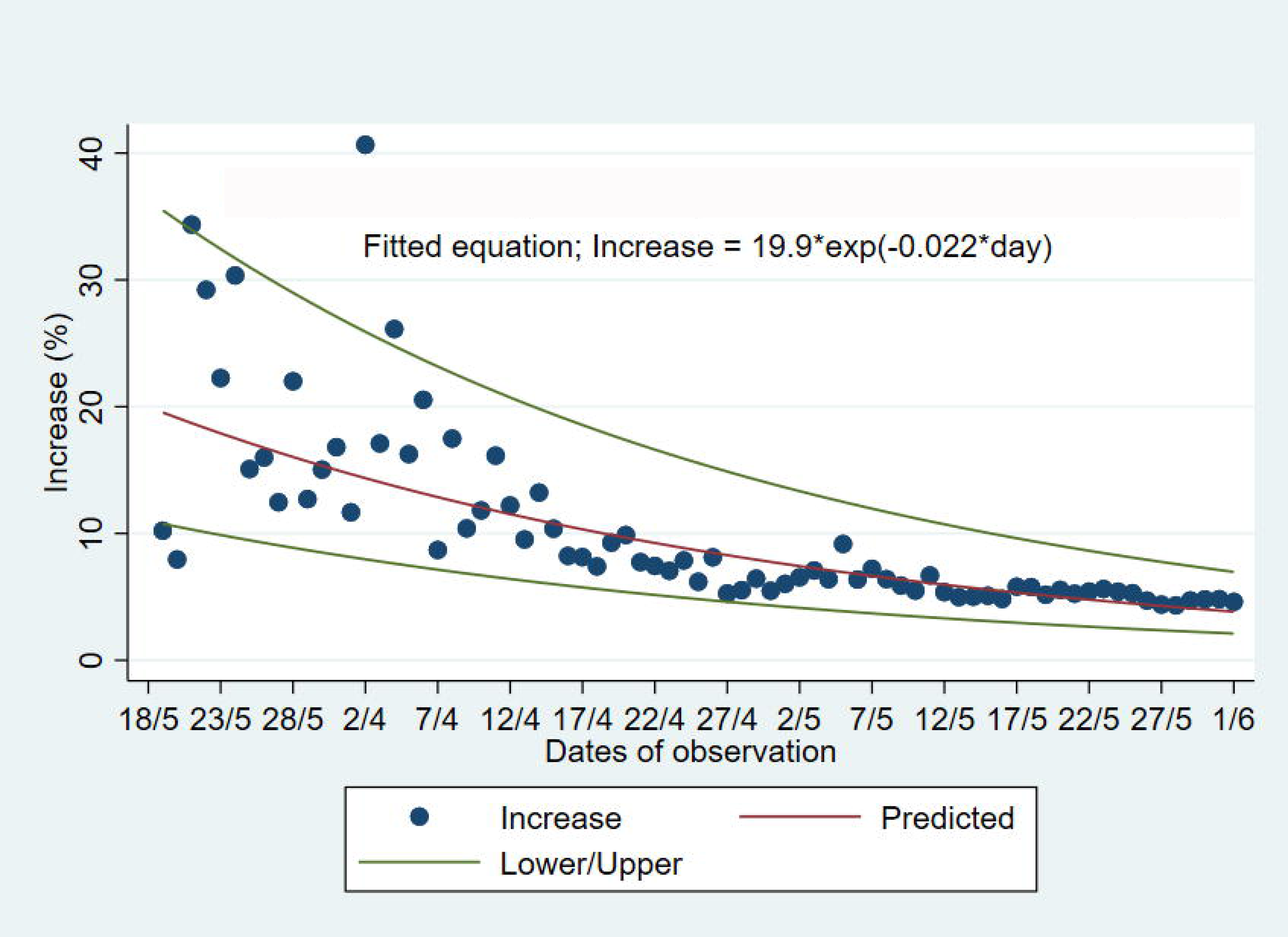
Exponential relationship between percent increase and time with 95% CI.

### Incidence rate per million population

During the study period, the incidence rate increased from 0.01 on March 19, 2020, to 6.14 on June 1, 2020. The average incidence rate with a 95% confidence interval (CI) was 1.84 (1.43-2.25). Day specific incidence rate revealed the exponential pattern, and the fitted equation is y= 0.062 * exp(0.069 * *t*), Where y-is daily incidence rate at time t-day, 0.062 is the early incidence rate per million population, and 0.069 is the instantaneous rate of increase per day. The fitted equation in terms of R^2^-adjusted value (0.886; P<0.001), AIC (121.9) and RMSE (0.54) were satisfactory.

### Doubling time of cases

Based on the instantaneous rate (0.069) of exponential growth curve we could estimate the doubling time of cases during the study period using the formula, days=ln(2)/r,*w*here ‘r’ is the instantaneous rate of increase. (0.069). Therefore, the doubling time with 95% CI was 10.05 (9.25-10.93) days. We also fitted the pattern of the cumulative number of cases recorded daily. We noted the pattern was similar to the incidence rate, and the fitted equation is y= 502.7 * exp(0.089 * *t*), Where y-is the cumulative number of cases on day ‘t’ and 0.089 is the instantaneous rate of increase. The R^2^-adjusted, AIC and RMSE values were 0.935, 114.4 and 0.512, respectively. The estimated doubling time with 95% CI was 7.79 (7.34-8.30).

### Case fatality rate (%)

Case fatality rate (CFR) varied between 1.41 and 3.43%. The overall CFR during the study period was 2.92% with 95% CI 2.82% −3.02%. As observed in the incidence rate, there was no clear trend in the CFR.

### The death rate per million population

The death rate during the study ranged from 0.002 to 3.948, and contributing the overall death rate was 1.14 (95% CI: 0.87-1.41) per million. There was a significant linear relationship between death rate (y) and incidence rate (x) with the estimated equation, y=0.050+0.029*x, (R^2^-adjusted value= 0.99; P<0.001; RMSE=0.080). The fitted equation suggested that for every 1000 new cases, the expected number of COVID-19 deaths would be 29 (95% CI: 28-30).

### Distribution pattern of new cases

Using day-specific new COVID-19 cases, we fitted three familiar distributions viz—Gamma, Weibull and Log-normal distributions using maximum likelihood estimation procedure. We presented parameters estimation and goodness fit statistics/criteria in Table 1. Of the three distributions fitted (Fig. 2), Gamma distribution revealed a good fit, as evidenced by the lowest goodness of fit statistics and goodness of fit criteria. Further, we evaluated the good fit of Gamma distribution by comparing the empirical density function, CDF, Q-Q plot (quantiles of data distribution), and P-P plot (empirical cumulative distribution function), and depicted in Fig. 3. We inferred that the Gamma distribution fairly represented with all empirical properties of the distribution and Fig.4 shows the empirical and theoretical cumulative distribution function (CDF) with 95% CI.

**Table 1:**
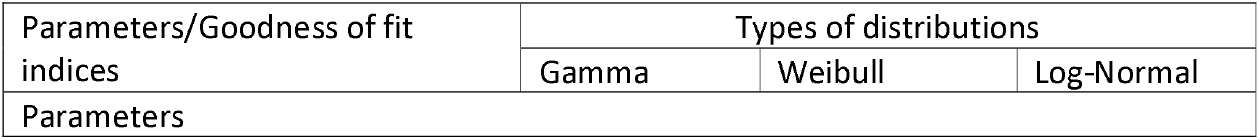
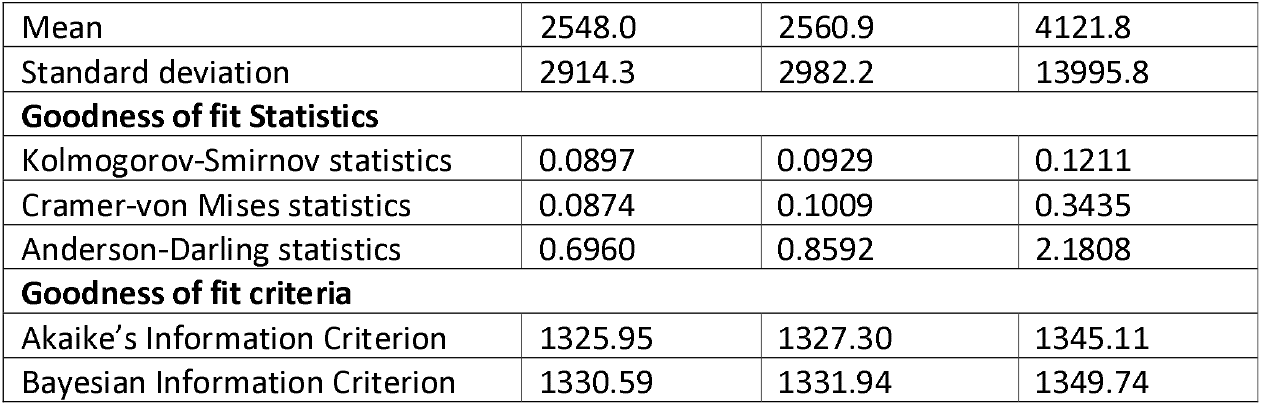
Parameters estimation and goodness of fit indices for three discrete distributions using maximum likelihood estimation.

**Fig 2.**
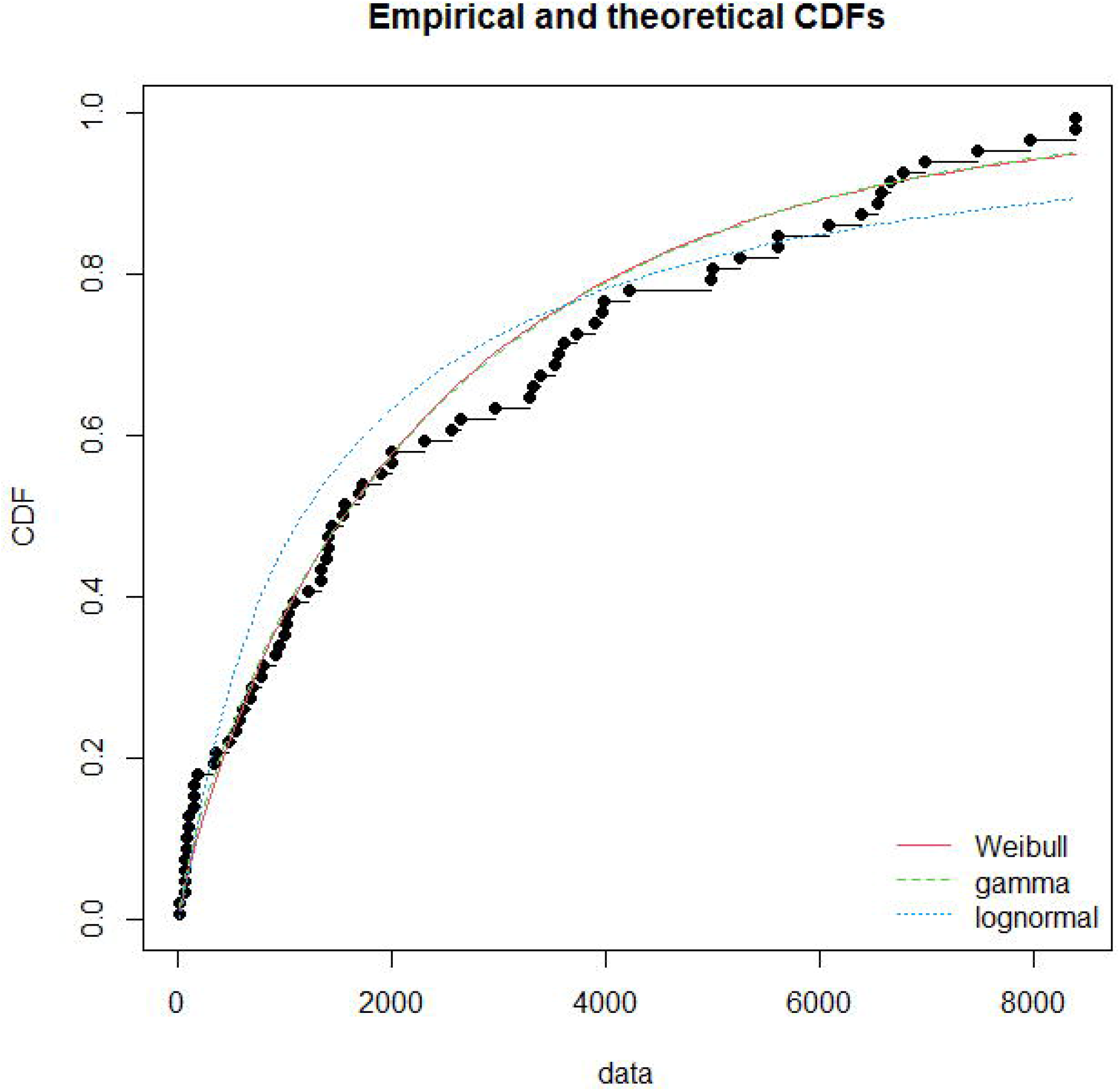
Statistical distributssions fitting with predicted values and empirical data.

**Fig 3.**
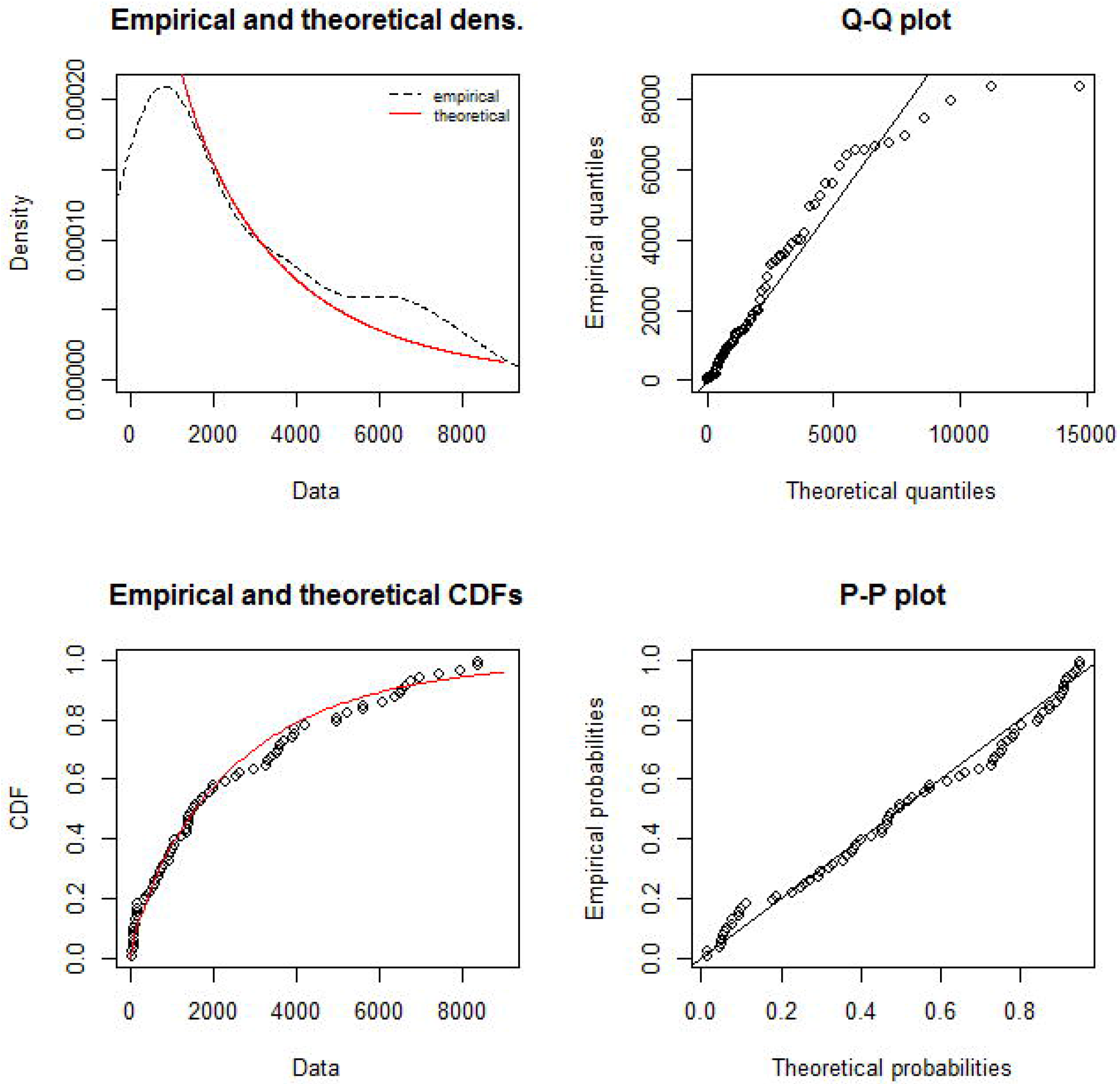
Good of fit assessment plots for Gamma distribution fitting.

**Fig 4.**
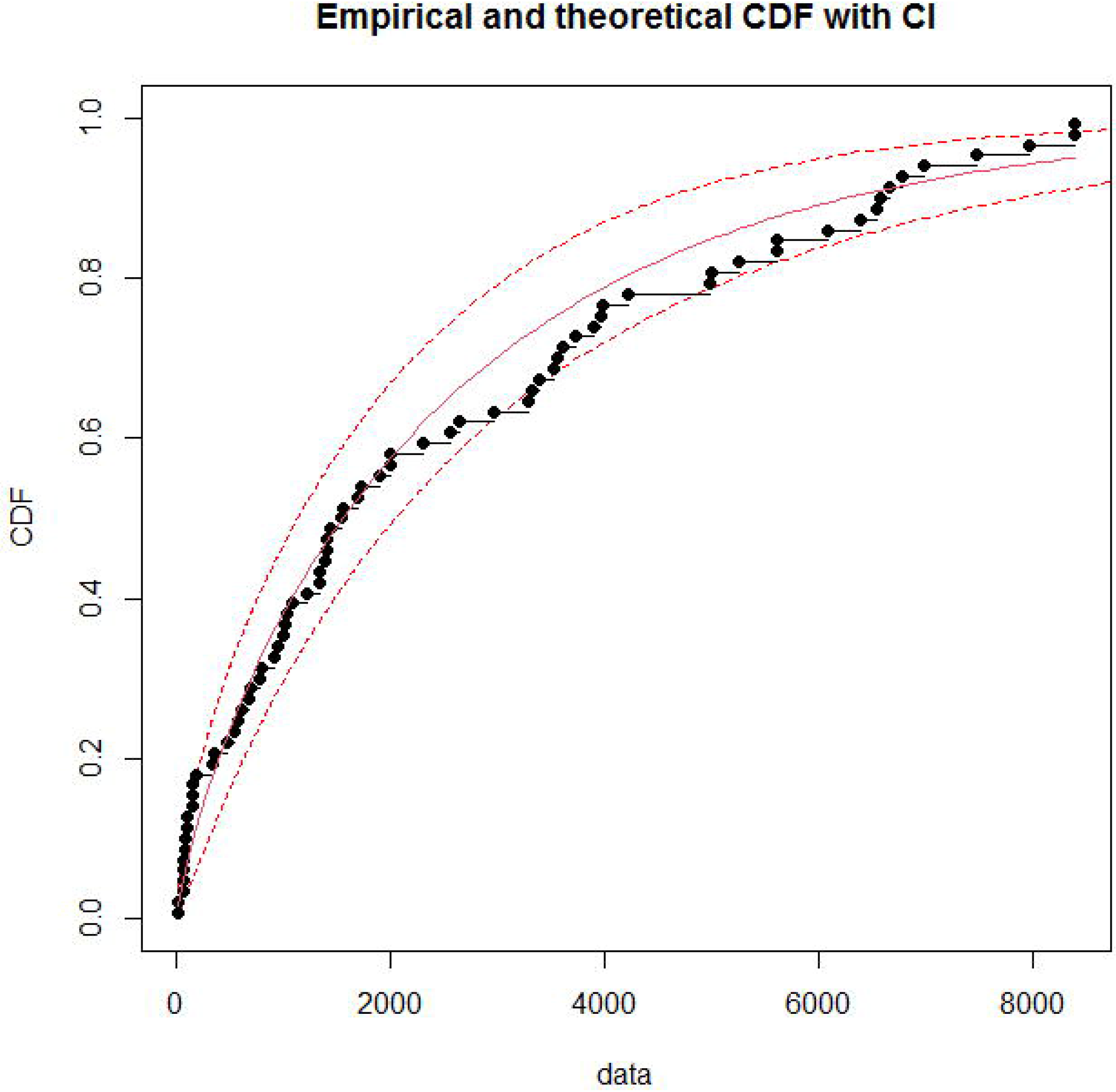
Gamma distribution fitting predicted values with 95% CI.

Since the Gamma distribution fitted well to the empirical data, to assess the consistency of estimating the parameters, we applied various estimation procedures such as MME, MSE and MGE. We presented parameters evaluation based on each process and goodness of fit criteria in Table 2. The parameters estimation by maximum likelihood method proved to be the best as shown by small values of the goodness of fit tests compared to the other procedures.

**Table 2:** Parameters (shape and rate) estimation of Gamma distribution by different methods.

| Parameters/Goodness of fit indices | Methods of estimation |  |  |  |
| --- | --- | --- | --- | --- |
|  | Maximum Likelihood estimation (MLE) | Matching Moment Estimation (MME) | Maximum Spacing Estimation (MSE) | Maximum Goodness of fit Estimation (MGE) |
| **Parameters** |  |  |  |  |
| Shape | 0.7644 | 1.0966 | 0.7353 | 0.6920 |
| Scale | 0.0003 | 0.0004 | 0.0003 | 0.0002 |
| **Goodness of fit criteria** |  |  |  |  |
| Akaike's Information Criterion | 1325.95 | 1333.28 | 1326.10 | 1327.15 |
| Bayesian Information Criterion | 1330.59 | 1337.92 | 1330.74 | 1331.78 |

### Generation Time (GT) estimation

We adopted four underlying distributions viz. empirical, Gamma, Weibull and Log-Normal for GT estimation. Among these four distributions, we could get a consistent estimate of GT (Table 3) for Gamma, Weibull and Log-normal. Since the Gamma distribution was the best fit for empirical data, we chose the GT based on the Gamma distribution as the reliable one and accordingly, the mean ± sd of GT was 14.2±11.9 days.

**Table 3:** Generation time estimation by different underlying distributions.

| Distribution | Mean generation Time (days) | Standard deviation |
| --- | --- | --- |
| Empirical | 56.4 | 14.81 |
| Gamma | 14.17 | 11.93 |
| Weibull | 14.26 | 11.93 |
| Log-normal | 13.95 | 11.68 |

### Estimation of Reproduction numbers (R0)

We presented R0 estimates based on the maximum likelihood estimate and exponential growth method for each phase of lockdown in Table 4. Since the incidence rate followed the exponential growth pattern, we considered the R0 estimate using the exponential growth pattern as a reliable estimate. While the R0 estimate during the pre-lockdown phase was 6.58 (95% CI: 5.02-8.39), it varied from 1.43 in the third phase to 3.19, in the first phase. During the four periods of the lockdown, the R0 estimate was 1.79 (95% CI: 1.78-1.80) suggesting a significant decline from the estimate of pre-lockdown phase. Further, because of the 70 days lockdown phase, the R0 estimate in the entire study period also was shown to be 1.83 (95% CI: 1.82-1.83).

**Table 4.** Reproduction number estimation based on different methods during the different phases of study period.

| Lockdown phases | Maximum likelihood estimation method |  | Exponential Growth method |  |
| --- | --- | --- | --- | --- |
|  | Reproduction Mean values | 95% confidence limits (LCL-UCL) | Reproduction Mean values | 95% confidence limits (LCL-UCL) |
| Lockdown phase 1 (25<sup>th</sup> March – 14<sup>th</sup> April ) | 3.55 | 3.45-3.65 | 3.19 | 3.11-3.28 |
| Lockdown phase 2 (15<sup>th</sup> April- 3<sup>rd</sup> May ) | 2.74 | 2.69-2.78 | 1.75 | 1.71-1.79 |
| Lockdown phase 3 (4<sup>th</sup> May - 17<sup>rd</sup> May ) | 3.25 | 3.21-3.29 | 1.43 | 1.39-1.46 |
| Lockdown phase 4 (18<sup>th</sup> May – 1<sup>st</sup> June) | 3.10 | 3.07-3.13 | 1.53 | 1.51-1.56 |
| During all the phases | 1.76 | 1.75-1.77 | 1.79 | 1.78-1.80 |
| Entire study period | 1.76 | 1.75-1.77 | 1.83 | 1.82-1.83 |

## Discussion

The merit of the present study is to understand the epidemiological situation of COVID-19 in India during the early transmission period, which covered the complete lockdown phase. Further, we presented various epidemiological indices based on empirical data, which will be more relevant for studying the transmission dynamics of the COVID-19 in a situation like a lockdown phase. This study findings also reflect the impact of the lockdown phase in India, which is ranked number two in the population size.

The aim of the lockdown during the early transmission phase was to delay the spike and to put health services and systems in place. The objective, to a large extent, has been met, as evidenced by our demonstration that the growth pattern of the cumulative number of cases was negative exponential. The observed percent increase in cases as on July 2, 2020 was also within the 95% CI prediction. Further, the data on COVID-19 maintained by our world in data (OWID) [6] showed that at the end of the lockdown phase, about 5% of total tested samples turned out to be positive. The similar figures on the same date in the United States, Mexico and the United Kingdom were 10%, 31% and 9% respectively. These facts highlight the impact of lockdown in India, which has a population density of about 450 persons per Km^2^. The effect is sustainable even during the post-lockdown phase as shown by only 6.3% of total tested found to be positive as on June 26, 2020 [6].

In many affected countries, the number of detected cases were considered as incidence in their respective countries. Since population size varies across countries, comparison of the cases between the countries is worthless unless it is standardized with the corresponding population size of the country. Therefore, in this study, we have considered the projected population (∼1366 million) in the year 2019 as a susceptible population, and it is very much pertinent for the second most populated country. While the average incidence rate per million people during the lockdown phase was about 2.0 in India, it was 75, 57 and 29 in the countries the USA, UK and France respectively in the same period as shown by OWID [6]. Further, the incidence rate (12.5) in India as on June 26, 2020, was significantly less than in the USA (123.7) on the same date.

We could estimate the doubling time, which explains the time taken in the community to get the doubling of cases. Based on the observed exponential growth curve on confirmed new cases as well as cumulative no. of cases, the estimated doubling time was 10 and 8 days. These estimates were higher than the estimates (1.4- 3.1 days) obtained in China [2,3,7]. Few more studies also reported that the doubling time was in the range of 6-7 days [8–12]. Higher doubling time is an indicator of a slowdown in the transmission during the lockdown phase. Case fatality rate [13,14] typically is used to measure the disease severity, and even though it is not an indicator for lockdown impact, it is essential information for policymakers to take appropriate action to prevent many deaths. Estimated CFR was 2.92% (95% CI: 2.82% - 3.02%) and the corresponding figures in USA and UK were 5.0% and 13.7% respectively as evidenced by OWID inferring that even disease severity is less in India compared to developed countries [13,15]. Average death rate 1.14 per million population in India was abysmally low compared to 161, and 323 observed in the USA and UK respectively. Further, the OWID data reveals that in India the percentage of people above 65 years age is 6% and the overall smoking prevalence is about 11.0%. In severely affected countries such as USA, UK, Spain, Italy, Germany, France and Belgium, the aged people (>=65 years) population varies between 18% (Belgium) and 23% (Italy). Similarly, the prevalence of smoking varies between 22% (USA) and 33% (France). These observations indicate that the aged people, coupled with high smoking status, may be a significant triggering factor for more COVID-19 deaths.

Understanding the distribution pattern of cases will be useful for estimating the vital epidemiological measures such as generation time and basic reproduction rate. Therefore, we fitted different statistical distributions to decide an adequate distribution of the observed data, and also we validated the estimations of the parameters by various methods. We obtained Gamma distribution as an adequate fit and established that the estimation of the parameters by maximum likelihood method as a reliable one. Confirmed cases provided the generation time (incubation period) estimation using different underlying distributions (Table 3). Though the estimates were consistent for the three statistical distributions, the generation time (14.2 ±11.9 days) using the Gamm distribution considered to be reliable. Estimations based on serial events [7,15–17], the generation time was higher in India, which implies that the majority of infected individuals might be typical asymptomatic during the early transmission stage. Another study ([18] estimated the median incubation period as 5.1 days (95% CI, 4.5 to 5.8 days). The authors further stated that 101 out of every 10 000 cases (99^th^ percentile, 482) would develop symptoms after 14 days of active monitoring or quarantine. These observations qualify our conservative estimate based on reported confirmed cases. Basic reproduction rate (R0) is a measure of transmission potential of any type of disease, and it is likely to vary because of the severity of the disease between the endemic countries. Since the outbreak of COVID-19, estimates using different approaches are available in various countries [7,10–12,17,19–22].

We could also estimate R0 using maximum likelihood and exponential growth methods during the different phases of lockdown. Our R0 estimates varied from 2.7 in the second phase to 3.6 in the first phase. On account of 70 days lockdown, the R0 estimate was 1.76 (95% CI: 1.75-1.77). During the lockdown phases 2-4, we observed that the R0 estimates obtained by exponential growth method were markedly less compared to the corresponding estimates obtained by MLE method indicating that the pattern of growth is restricted due to lockdown phase. The R0 estimate in different countries [21] varied from 1.6 in Republic Korea to 8.2 in the USA. Further, in a meta-analysis study [20] conducted based on various studies carried out in China had shown that the R0 estimate by exponential growth method varied between 1.9 and 6.3. Compared to these observations, our overall estimate (1.79; 95% CI: 1.78-1.8) was much less than the pooled estimate of R0 (3.32; 95% CI: 2.81-3.82) obtained in the meta-analysis.

Several studies [23–27] on COVID-19 had estimated various transmission/epidemiological indices in different epidemiological situations using mathematical modellings approach. Indeed, the models will unravel the estimates of many parameters, which may not be measured directly. Further, the models will be used to forecast the disease outbreak to take appropriate preventive measures. However, the accuracy of the model prediction depends on the input parameters that reflect the real epidemiological situation. Therefore, in this context as a principal analysis, we carried out a real data-based analysis to assess various epidemiological measures. Further, these measures area assumptions free estimates in an early stage of disease transmission, which was under lockdown pressure.

## Data Availability

All data are available in our world in data (OWID) public domain as well as in WHO Corona situation reports.

https://github.com/owid

## Acknowledgement

Ministry of Health and Family Welfare, Government of India (GOI), WHO and OWID are grately acknowledged for accessing the public domain data.

## Author’s contribution

PV-formulated the study design, generated database, analysed and wrote the manuscript.

